# Are we #Stayinghome to Flatten the Curve?

**DOI:** 10.1101/2020.05.23.20111211

**Authors:** James Sears, J. Miguel Villas-Boas, Vasco Villas-Boas, Sofia Berto Villas-Boas

**Affiliations:** Department of Agricultural & Resource Economics; University of California, Berkeley; Haas School of Business; University of California, Berkeley; Department of Economics and Department of Electric Engineering and Computer Sciences; University of California, Berkeley

## Abstract

The recent spread of COVID-19 across the U.S. led to concerted efforts by states to flatten the curve through the adoption of stay-at-home mandates that encourage individuals to reduce travel and maintain social distance. Combining data on changes in travel activity with COVID-19 health outcomes and state policy adoption timing, we characterize nationwide changes in mobility patterns, isolate the portion attributable to statewide mandates, and link these reductions to changes in COVID-19 health outcomes. We find evidence of dramatic nationwide declines in mobility prior to adoption of any statewide mandates. Once states adopt a mandate, we estimate further mandate-induced declines between 2.1 and 7.0 percentage points relative to pre-COVID-19 baseline levels. Controlling for pre-treatment changes in mobility, we estimate mandate-induced declines of 0.13-0.17 fewer deaths and 5.6-6 fewer hospitalizations per 100,000 residents, equivalent to 23-30,000 averted deaths in adopting states for the months of March and April and death rates 42-54% lower than in the absence of policies. These estimates represent a likely lower bound on the health impacts of stay-at-home mandates, and our findings for the health impacts of early mobility reductions convey important policy implications for re-opening.

## Introduction

Since December 2019, the novel coronavirus SARS-CoV-2 (COVID-19) has spread rapidly around the world and in the U.S., prompting dramatic policy responses. Local, state, and national governments around the world have an extensive set of policy instruments with which to fight the pandemic and limit the virus’ impact on their constituents. Because many regions have exhibited exponential growth in coronavirus cases, policymakers are increasingly implementing aggressive stay-at-home mandates to reduce transmission through human interaction in order to “flatten the curve” [28]. As of March 31, the U.S. had the highest number of confirmed cases (more than 67% more than the next country) with at least one resident of every state affected [35]. Improving our understanding of how existing stay-at-home policies reduce travel activity and ultimately mitigate negative health consequences of the pandemic will help local and state policymakers determine the optimal policies to help “flatten the curve” and quell the spread of COVID-19. To investigate this, we combine data on human mobility with state policy variation and health outcomes, allowing us to determine the reductions in distance traveled, visits to non-essential businesses, and human encounters, and ultimately relate these to changes in hospitalizations and deaths.

The relevant benefits of non-pharmaceutical interventions (NPI) – such as quarantining infected households, closing schools, and banning social events or large gatherings - have largely been informed by mathematical models [22]. In addition, some anecdotal and historical evidence supports their efficacy. In California’s San Francisco Bay Area, the first area of the country to implement stay-at-home mandates, doctors reported “fewer cases than expected” after two weeks of social distancing [25]. Analysis of internet-connected thermometers suggests that new fever rates on March 23 were below those at the start of the month, while state hospitalization rates showed a commensurate decline in growth rates. Washington state officials reported similar reductions in COVID-19 transmission as a result of the state’s containment strategies [5]. Exploration of death rates and NPI rollout in 17 U.S. cities during the 1918 influenza pandemic support these claims, finding that implementation of multiple social distancing practices intended to reduce infectious contacts early in the outbreak led to 50% lower peak death rates and flatter epidemic curves relative to cities that did not implement such policies [22]. Gaining insight into the effectiveness of these stay-at-home mandates is critical for understanding the benefit of making the considerable economic sacrifices required to enact such policies. Even before mandates limited economic activity, GDP forecasts suggested an economic contraction in the U.S. of 24% [27]. Concerns over these costs prompted comments from the executive branch regarding relaxation of restrictions and allowing non-essential businesses to reopen, prompting opposition from public health experts [18] and many economists [23].

Recent simulations provide further insight into the benefits of social distancing. While epidemiological models of the U.K. and U.S. suggest that techniques for mitigating exposure of those most at risk may drastically reduce peak load on the healthcare system and cut COVID-19 deaths by half, such techniques on their own might not be enough to prevent the healthcare system from being overwhelmed. Some argue that, in this case, a combination of social distancing, self-quarantine of infected people, and suspension of schools would need to be maintained until a vaccine is available to prevent a rebound [17]. Other experts call for widespread testing coupled with digital contact tracing as a means to reduce viral spread while minimizing harmful social and economic side-effects. Simulations based on a moderate mitigation policy (comprising 7-day isolation following any symptoms, a 14-day quarantine for the household, and social distancing for all citizens over age 70) find that, had it been implemented in late March, it would have reduced potential U.S. deaths by 1.76 million [21]. Given that this simulated policy is less stringent and maintained for a shorter duration than many of the policies currently observed, the actual benefits from existing stay-at-home mandates (either directly from reduced COVID-19 deaths or indirectly due to decreased transmission of other illnesses) from existing stay-at-home mandates could be substantially larger.

This paper contributes to the existing literature first by looking at mobility patterns during the pandemic across states and time and second by providing the first empirical evidence of stay-at-home policies’ effectiveness. These mandates combine closures of non-essential businesses with instructions for all residents to remain at home except for the purchase of necessities (i.e. groceries or medicine), with the goal of limiting “unnecessary person-to-person contact” [29] and to “mitigate the impact of COVID-19” [10]. First, we examine changes in travel behavior across the United States in response to these social distancing policies and estimate the portion of these reductions attributable to early state stay-at-home mandates. Then, we examine changes in health outcomes and estimate the portion of these changes attributable to state stay-at-home mandates, while controlling for other potential factors.

To estimate the changes in travel activity and social distancing since the spread of COVID-19 in the United States, we use data on changes in average distance traveled, visits to non-essential businesses, and unique human encounters per square kilometer by day and by state [33] relative to pre-COVID-19 baseline levels. Through data visualization and descriptive event studies, we show that tremendous nationwide reductions in travel activity levels occurred prior to statewide mandates, suggesting residents were already responding to local policies and perceived risks. Prior to any state implementing a statewide mandate, average travel distances had already fallen by 16 percentage points, the human encounter rate by 63 percentage points, and non-essential visits by 39 percentage points relative to pre-COVID-19 levels, providing evidence of extensive social distancing occurring even before statewide orders requiring such behavior.

We then estimate a differences-in-differences model that isolates the effect of statewide mandates by comparing differences before and after mandate implementation and between early-adopting, later-adopting, and control states. Using this framework, we test whether states’ stay-at-home policies induced significant changes in mobility and daily human encounters in the United States once mandates were implemented. After presenting results from the difference-in-differences model, we discuss estimates from event study models that directly examine the dynamic effects of stay-at-home mandates. In addition to typical unweighted event studies, we implement weighted event studies that directly balance on differences in pre-adoption outcome trends [6]. Results suggest similar effects as the difference-in-differences estimates, supporting the finding that statewide mandates induced further reductions in travel activity even after considerable pre-mandate reductions.

Across both methods, we find evidence that residents reduced daily activity, even before mandates, and that patterns differ by state. Moreover, we estimate significant additional reductions in travel and increased social distancing in response to stay-at-home mandates. We estimate a 7.0 percentage point reduction in average distance traveled, a 2.1 percentage point decline in non-essential visits, and a 3.5 percentage point reduction in the daily rate of human encounters after the average stay-at-home mandate was implemented.

Using comparable difference-in-differences and event study methods, we test whether states’ stay-at-home policies induce significant changes in COVID-19 health outcomes in the United States. We find evidence that stay-at-home mandate adoption is associated with a decline in daily death rates of 0.13-0.16 fewer deaths and 5.6-6.0 fewer hospitalizations per 100,000 population. Moreover, we find that changes in non-essential visits in the pre-mandate period further contributed to reducing death rates. We estimate that stay-at-home mandate-induced behavioral responses likely averted 23-30,000 COVID-19 deaths during observed mandates in the months of March and April. Adding in averted deaths due to pre-mandate social distancing behavior, we estimate a total of 48-71,000 averted deaths from COVID-19 for the two-month period. Given that the actual COVID-19 death toll for March and April was 55,922, our estimates suggest that deaths would have been 1.86-2.27 times what they were absent any stay-at-home mandates during this period.

Taken together our results provide evidence that the implementation of non-pharmaceutical interventions in the form of statewide stay-at-home mandates encouraged additional social distancing and likely helped further reduce the spread of the pandemic. To our knowledge, ours is the first paper that investigates mobility during the COVID-19 pandemic and provides evidence of reduced travel activity, increased social distancing, and of associated health benefits resulting from stay-at-home mandates and pre-mandate mobility declines. We contribute to the overall understanding of the direct health benefits of current COVID-19 policies and provide evidence that these policies are having the intended effect of reducing social interactions and reducing negative health consequences from the current pandemic. Our findings of substantial reductions in mobility prior to state-level policies convey important policy implications.

## Materials and Methods

### Mobility Data

We obtain travel activity and social distancing data from the analytics company Unacast [33]. To understand how well different communities are social distancing, Unacast uses cellular location data for 15-17 million identifiers per day to construct three measures of behavior in response to COVID-19 policies. Each measure is aggregated to the state-by-day level and is defined as the daily percentage point change relative to that weekday’s average for the pre-COVID-19 period of February 10 through March 8 (henceforth referred to as the baseline period). While all data are published directly to their Social Distancing Dashboard in the form of figures and maps [33], we obtained the balanced panel of state-by-day observations for the period of February 24 through April 29, 2020 directly from Unacast. Unacast receives location data from millions of mobile devices through authorized applications, Wi-Fi or Bluetooth connections, and A-GPS positions. Obtained information includes the location of the device at a given point in time (latitude, longitude, and elevation) along with the mobile device make, model, and operating system, the corresponding application gathering the data, GPS accuracy value, and the direction and rate of travel. Each state-day observation we use is calculated using position information. Taken together, these three measures paint a comprehensive picture of behavior changes in response to states’ stay-at-home mandates. See Appendix A for more details on the data collection process, the equations used to construct each measure, and further discussion on sample composition and potential biases or measurement errors.

The first metric we use is the change in average distances traveled (*AḊT*), which provides a measure of overall changes in travel activity during the COVID-19 period. The average distance traveled across all devices assigned to a state on a given day is compared to the state’s average for that day of the week during the pre-COVID-19 baseline period. As a result, *AḊT* measures the percentage point change in average distance traveled relative to more typical behavior prior to the pandemic, accounting for pre-existing differences in states’ average mobility patterns throughout the week.

Changes to average distances traveled give a sense of broader transformations to travel behavior. Declines in relative activity will yield negative values of *AḊT*, which can reflect both intensive margin (short distances traveled for the same frequency of trips) and extensive margin (some trips foregone entirely) adjustments. Reductions in *ADT* following mandate implementation would reflect compliance on average with states’ guidances to work from and stay at home except for essential activities. A value of *AḊT_it_ =* 0 indicates that the average distance traveled for individuals assigned to state *i* on date *t* was identical to the pre-COVID-19 distance for that day of the week. A value of −7 conveys that, on average, devices assigned to the state traveled an average distance 7 percentage points less than during the pre-COVID-19 baseline. This approach allows us to account for patterns in propensity to travel throughout the week, making sure our comparison accurately reflects the previous average conditions for a particular day of the week.

The second metric we use is the change in visits to non-essential businesses, defined as Non-Essential Visits (*NĖV*). To the extent that non-essential businesses closed following stay-at-home mandates, we expect to see reductions in the number of trips residents take to these types of retail or service businesses. Our utilized measure of the change in visits to non-essential businesses (*ṄEV*) offers a similar comparison to *AḊT* that is targeted at travel to the types of businesses most heavily impacted by stay-at-home mandates. Businesses likely to be deemed “non-essential” include department stores, spas and salons, fitness facilities, event spaces, and many others. To improve accuracy, non-essential businesses are defined according to group definitions in both the Unacast SDK and the OpenStreetMaps POI’s.^1^ *ṄEV* is calculated by dividing a state’s average number of visits to non-essential businesses by its day-of-week baseline level. A value of *NĖV_it_ =* 2 indicates a two percentage point increase in visitations to non-essential businesses relative to baseline norms for that weekday in a given state.

Finally, we use changes in the rate of unique human encounters (*EṄC*) as a measure of social distancing. While *AḊT* and *NĖV* provide information on two potential margins for adjusting travel behavior, neither directly captures changes in human-to-human interaction. As COVID-19 transmission primarily occurs through “close contact from person-to-person,” having a measure of potential human encounters allows us to further understand whether reductions in travel distance and business visitations translate into fewer opportunities for transmission [13]. *EṄC* measures the change in the rate of unique human encounters per square kilometer relative to the state’s baseline levels. Following [30], one unique encounter is produced every time two devices assigned to a given state are observed within a 50 meter radius circle of each other for no more than 60 minutes.^2^ Dividing the state-level sum of encounters for the day by the state’s square kilometer of land area provides the state’s daily rate of unique human encounters. Finally, this encounter rate is normalized by the state’s average encounter rate for that day of the week during the baseline period.^3^ As a result, an encounter rate equal to that of the state baseline rate for that day of the week results in a value of *ENC_it_ =* 0, while a value of *ENC_it_ =* −12 indicates a 12 percentage point reduction in the encounter rate for state *i* on date *t* relative to the state’s pre-COVID-19 level for that day of the week.

Table 1 provides summary statistics for the three mobility measures across the sample period. The table is divided into four panels. The first panel corresponds to February 24-March 8.^4^ The second panel summarizes behavioral changes for the remainder the month of March, and the third panel for April 1 to April 29, 2020. The final panel provides the average, median, and standard deviation for the total sample. The three columns report summary statistics for the change in average distance traveled (*AḊT*), non-essential visits (*NĖV*), and the unique human encounter rate (*EṄC*).

**Table 1:**
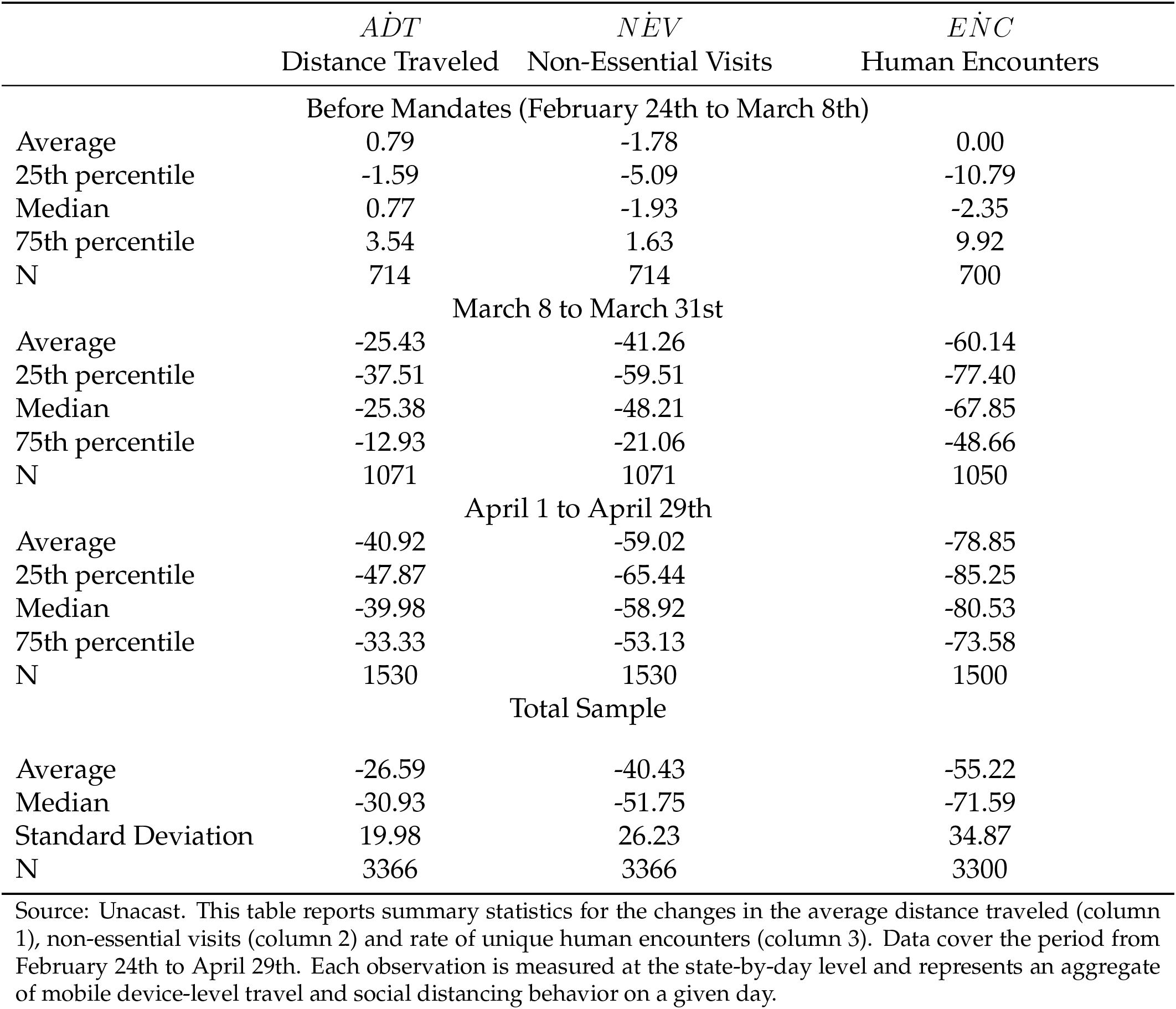
Summary Statistics on Travel Behavior and Social Distancing

In the top panel, covering the end of February until March 8, we see that the daily average distance traveled was larger than pre-COVID-19 baseline levels by 0.79 percentage points. In column 2, we see that non-essential visits were 1.78 percentage points lower than the pre-COVID-19 baseline, with the rate of human encounters on average at baseline in column 3.^5^ For the month of March, all three mobility measures experience large decreases relative to pre-COVID-19 levels, attesting to average reductions in both travel and social interactions. In the month of March, we observe 75th percentile changes of 12.93 percentage point reductions in average distances traveled, reductions for non-essential visits of 21.06, and encounter rates 48.66 percentage points below baseline levels. Travel reductions become even more dramatic in the month of April; average reductions in all three travel activity measures during April exceed in magnitude the equivalent March declines.

Our utilized travel measures display very high correlations with travel data produced by other sources. Comparing our utilized travel change variables with the measure of state-level changes in retail and recreation travel from Google’s COVID-19 Community Mobility Reports, we find average correlations of 0.95 with *AḊT* and 0.98 with *NĖV* [20]. Changes in non-essential visits are nearly perfectly correlated with the Google measure, with no state exhibiting a correlation below 0.96. Correlations for *AḊT* remain high but exhibit greater variation across states. 31 states have correlations above 0.95, including California and New York at 0.97 and 0.98, respectively. Wyoming displays the lowest correlation at 0.75. These strong relationships across data providers suggest that our results are indicative of general mobility patterns and not spurious results arising from anomalies of our chosen data source.^6^

### Stay-at-Home Mandate Data

To denote periods before or after a state implemented a “stay at home order,” we obtain the date each statewide policy was issued for all fifty states and the District of Columbia [28]. We define our early adopters as the first four states to pass a stay-at-home mandate: California (March 19), Illinois, New Jersey, and New York. The second group comprises the 38 late adoption states and D.C.. The last group comprises the eight remaining states that never implemented statewide mandates: Arkansas, Iowa, Nebraska, North Dakota, Oklahoma, South Dakota, Utah, and Wyoming. The observed stay-at-home mandates all consisted of a mix of specific non-pharmaceutical interventions; each observed policy closed or placed considerable limits on non-essential businesses and required residents to stay at home except for essential activities. Essential services include grocery stores, gas stations, pharmacies, banks, laundry services, and business essential to government functions [11]. Throughout this paper, we refer to all mandates that implement this combination of policies as a “stay-at-home mandate.”

While we focus our attention on statewide stay-at-home policies, many county and local policies had already been implemented and were already affecting individual-level mobility. Six San Francisco Bay Area counties required residents to stay-at-home beginning March 17, two days prior to the statewide mandate. By mid-March, schools of all levels had begun closing their doors and transitioning to online instruction. On March 9, Stanford University moved classes online “to the extent possible,” with Harvard and many other institutions swiftly following suit [24]. Further, business leaders including Google, Microsoft, Twitter, Facebook, and Amazon transitioned some or all of their employees to working remotely well before statewide mandates entered into effect [4]. As a result, any behavioral response to statewide stay-at-home mandates represents only a partial response to the suite of actions and policies undertaken to combat the spread of COVID-19. Our estimated “mandate effects” that follow therefore capture the behavioral responses specific to statewide stay-at-home mandates and underestimate the effect of all combined policies. If local, county, and business policies had already incentivized residents to stay at home, then we would expect a reduced response to later statewide mandates (which would be reflected in small magnitude estimates in our models). As a result, all estimated mandate effects that follow reflect mobility responses in addition to those already realized by pre-existing policies.

### Health Outcome Data

We obtain information on hospitalizations and deaths due to COVID-19 by state from the COVID Tracking Project (CVT) [15]. CVT obtains data on positive and negative tests, deaths, hospitalizations, and the counts of patients currently in intensive care units and on ventilators. Outcome data are obtained directly from the respective public health authorities and are supplemented with additions from press conferences or trusted news sources when applicable. While states consistently report both daily changes and total counts for COVID-19 deaths, hospitalization data are much more sparse and reported in different ways by different states. As a result, we are able to construct daily changes in deaths for every state on all sampled days (*N* = 3, 366), but on roughly one-third of days for hospitalizations (*N* =1,122). In total, 28 of 43 adopting states and 7 of 8 never-mandate states report hospitalization numbers at any point during the sample period (with NY the only early adopter with hospitalization data). As the bulk of data are obtained directly from state public health bodies, CVT represents one of the most transparent and up-to-date source of COVID-19 mortality and morbidity data. CVT counts of COVID-19 mortality correlate strongly with mortality measures obtained from other sources^7^ We scale the change in hospitalizations and deaths by one hundred thousand state population (using state population from the 2010 U.S. Decennial Census [34]).

Health outcome summary statistics are provided in Table 2. Summary statistics for COVID-19 mortality and morbidity for mandate and never-mandate states are presented in four panels. The first panel provides summary statistics for the observed baseline period (February 24 through March 8) while the second panel uses data for the entire pre-mandate period of February 24 through March 18. The third panel provides equivalent summaries for the mandate adoption period of March 19 through April 8, while the bottom panel covers the post-adoption period of April 9 through April 29. Hospitalization data is only obtained for one state (AZ) prior to March 19, resulting in missing or near-zero values in the top two panels.

**Table 2:** COVID-19 Mortality and Morbidity Summary Statistics

|  | Mandate States and D.C. | Never-Mandate States |
| --- | --- | --- |
| <u>Observed Baseline (Feb 24 - Mar 8)</u> |  |  |
| Mean Hosp. Rate per 100K | 0.0078 | NA |
| Mean Death Rate per 100K | 0.0008 | 0 |
| Median Death Rate per 100K | 0 | 0 |
| 25th Percentile Death Rate | 0 | 0 |
| 75th Percentile Death Rate | 0 | 0 |
| Average State Cumulative Deaths | 0.7209 | 0 |
| Median State Cumulative Deaths | 0 | 0 |
| Mean Days with Positive Deaths | 0.2558 | 0 |
| <u>Before Mandates (Feb 24 - Mar 18)</u> |  |  |
| Mean Hosp. Rate per 100K | 0.0704 | NA |
| Mean Death Rate per 100K | 0.0019 | 0.0006 |
| Median Death Rate per 100K | 0 | 0 |
| 25th Percentile Death Rate | 0 | 0 |
| 75th Percentile Death Rate | 0 | 0 |
| Average State Cumulative Deaths | 3.5349 | 0.1250 |
| Median State Cumulative Deaths | 0 | 0 |
| Mean Days with Positive Deaths | 1.4186 | 0.1250 |
| <u>Adoption Period (Mar 19 - Apr 8)</u> |  |  |
| Mean Hosp. Rate per 100K | 1.1031 | 0.3518 |
| Mean Death Rate per 100K | 0.1859 | 0.0354 |
| Median Death Rate per 100K | 0.0560 | 0 |
| 25th Percentile Death Rate | 0 | 0 |
| 75th Percentile Death Rate | 0.1662 | 0.0537 |
| Average State Cumulative Deaths | 352.3023 | 19.7500 |
| Median State Cumulative Deaths | 74.0000 | 12.5000 |
| Mean Days with Positive Deaths | 15.2093 | 7.8750 |
| <u>Post-Adoption (Apr 9 - Apr 29)</u> |  |  |
| Mean Hosp. Rate per 100K | 1.2475 | 0.3709 |
| Mean Death Rate per 100K | 0.5700 | 0.0974 |
| Median Death Rate per 100K | 0.2349 | 0.0686 |
| 25th Percentile Death Rate | 0.1046 | 0 |
| 75th Percentile Death Rate | 0.6306 | 0.1487 |
| Average State Cumulative Deaths | 932 | 50 |
| Median State Cumulative Deaths | 224 | 37 |
| Mean Days with Positive Deaths | 18.7 | 14.0 |

Source: The COVID Tracking Project (CVT). This table presents summary statistics for data on mortality and morbidity due to COVID-19 for mandate and non-mandate states across four periods. The top panel reports summary statistics for the observed baseline period of February 24 through March 8, while subsequent panels report statistics for the entire pre-mandate period of February 24 through March 18, the staggered adoption period of March 19 through April 8, and the post-adoption period of April 9 - 29. COVID-19 death and hospitalization rates are measured per 100 thousand population. N = 3,366 for death data and N = 1,122 for hospitalization data.

Across the country, very few deaths occurred prior to the adoption of statewide mandates. During the observed baseline period (top panel), only Washington state had experienced deaths resulting from COVID-19 with the state’s first death recorded on February 26. Through March 18 (second panel) 22 states had experienced positive deaths, with mean death totals of 3.5 for mandate states and 0.13 for non-mandate states (South Dakota reported one death on March 18).

Hospitalization and death rates rise considerably during the adoption period (third panel) with considerable state-to-state variation, reflecting the tremendous, heterogeneous spread of COVID-19 throughout the country during this period. While mandate states averaged 1.10 hospitalization and 0.19 deaths per 100,000 population from March 19 through April 8, over 25% of state-day observations saw no deaths. On average, mandate states reported over 352 total deaths – well above the median total of 74 – and positive death counts for over 15 of 21 days in the period. Never-mandate states exhibit markedly lower COVID-19 mortality and morbidity, averaging under 20 total state deaths with mean death rates over five times lower than mandate states (0.035) and more than half the mean hospitalization rate (0.35).

Turning to the post-adoption period (bottom panel), hospitalization rates plateau while death rates continue to rise. From April 9 to April 29, the average hospitalization rates per 100,000 population rose slightly to 1.25 for mandate states and 0.37 for non-mandate states while death rates nearly tripled in non-mandate states (to 0.0974) and more than tripled in mandate states (to 0.57). While average death counts for mandate states had reached 932, the median count of 224 and spread between the first quartile (0.10) and third quartile (0.63) death rates convey the tremendous variation in COVID-19 mortality even within states that chose to adopt stay-at-home policies. Death counts and rates remain much lower for never-mandate states, with mean, first quartile, median, and third quartile death rates all one-fourth or less of those for mandate states – indicating tremendous disparities in COVID-19 mortality across the two groups.

While our measures of mortality and morbidity represent the most up-to-date data available, they are still preliminary and likely represent underestimates of the true impacts of COVID-19. Widespread lack of access to testing early on in the U.S. outbreak and especially in rural areas means many deaths (especially at-home deaths) caused by COVID-19 may have gone uncounted [9]. As more information becomes known, state health authorities are likely to update their reported counts. Because CVT actively updates their data and we pull data directly from CVT each day, the numbers used in this paper accurately reflect mortality and morbidity information as currently known at the time of writing. The true public health impact of COVID-19 will likely not be known for years to come as reporting protocol is improved and prior deaths are verified.

### Empirical Strategy

#### Difference-in-Differences Model under Staggered Adoption

To determine the effect of statewide stay-at-home mandates on the outcome of interest, we begin by estimating the following difference-in-differences model under staggered adoption:

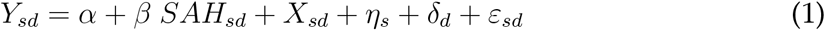

The outcome *Y_sd_* denotes the change in a given measure of travel activity (*AḊT*, *NĖV*, *ĖNC*) for state *s* on date *d* relative to the baseline level for mobility analyses, and either the death rate or hospitalization rate per 100,000 population for health analyses. Each outcome is expressed as a function of a constant *α*, whether a state has a statewide mandate in effect, and both time and unit fixed effects. *SAH* is an indicator equal to one if state *s* has a stay-at-home mandate in place on date *d* and zero otherwise. In the sections that follow, we consider the two cases where *SAH* includes variation for just the first four states to adopt statewide mandates and for all states that ever adopted a statewide mandate. The vector of state fixed effects *η_s_* controls for time-invariant characteristics of states that affect the outcome, while date fixed effects *δ_d_* control for factors affecting the outcome on a given date common to all states (i.e. executive branch press conferences or daily changes in worldwide total deaths/hospitalizations). The term *ε* is an idiosyncratic error comprised of unobserved determinants of changes in the outcome that are not controlled for by the variables specified in the linear Eq 1. Rather than include state-specific time trends, our preferred specifications use outcomes residualized of timing cohort pre-trends [19]. For health analyses we include a vector of time-varying controls *X_sd_* that account for average changes to mobility patterns in the pre-mandate periods for each of our three metrics.

The coefficient *β* measures the difference in the change in average outcome for states that have implemented a stay-at-home mandate relative to the change in activity in states that had yet to implement or never implemented such policies, after controlling for state and time-varying factors that also correlate with the outcome of interest. In this way 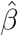 provides an estimate of the average treatment effect for treated states (ATT). Models for mobility variables are estimated using data for the entire sample period of February 24 through April 29. For health analyses we report estimates for the entire sample period as well as for two subsamples: pre-period observations trimmed to March 19 for never-mandate states and two weeks prior to adoption for mandate states (denoted *2wk Pre*), and excluding all observations for never-mandate states (*SAH* sample).

This empirical approach allows us to identify the relationship between stay-at-home mandates and daily changes in each of the ouitcomes of interest while also explicitly controlling for other confounding factors that are specific to each state or date. The shares of local population previously working from home or employed are controlled for with *η*, while day-to-day changes in factors common to all states – motivated by new information on the virus’ spread and nationwide media coverage or federal appeals for social distancing – are controlled for through *δ*. The mandate effect *β* is identified under the assumption that, after controlling for cohort-specific pretrends, common day-to-day trends, and time-invariant state characteristics, stay-at-home mandates are as good as random. Equivalently, the day-to-day outcome changes in states that had yet to adopt or never adopted a mandate are what the change in the outcome would have been for stay-at-home states absent the mandate. Given the time-varying nature of adoption, we can express this underlying assumption as the weighted average of parallel trends for each simple two-by-two DD estimators [19]. Our approach is identified using changes in the outcome that differ from typical pre-COVID-19 levels for a state and the states average change during the COVID-19 time. A remaining source of bias would be if the early mandate states were trending differently than the control states before March 18 in ways that differed from trends after March 18, or if similar differences in trends exist across all mandate states. Standard errors of the estimated parameters are clustered by state to account for variation in state policies potentially affecting the magnitude of the error term *ε*.

#### Unweighted and Weighted Event Studies

To directly model the dynamic nature of mobility and health responses to statewide stay-at-home mandates, we employ two event study methods. First, we estimate traditional event studies equivalent to the preferred difference-in-differences specifications:

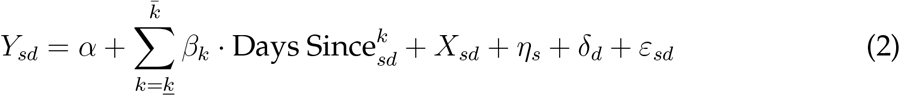

where

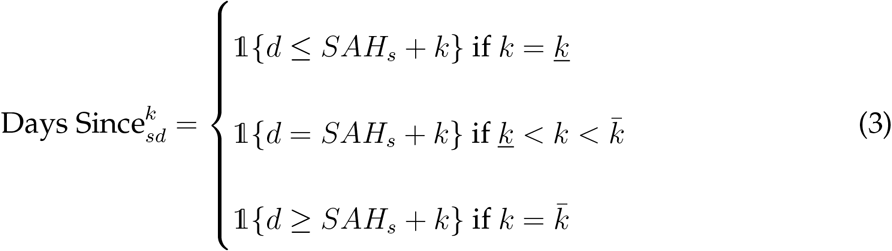

In this fashion the difference-in-differences estimator is decomposed into 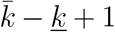 individual coefficients relating the impact of being *k* days relative to when a state adopted its statewide mandate on date *SAH_s_*. Event-time effects are identified under identical conditions as the staggered difference in differences model of Eq. 1. To ensure that dynamic treatment effects are identified separately from time trends even in models that omit never-treated states, the endpoints 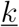 and 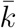 are binned to include all dates that fall either before 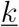 or after 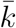 [31]. Following convention, we drop the *k* = − 1 bin and normalize all event-time effects relative to the day prior to mandate adoption.

Second, we estimate weighted event studies to further account for differences in pre-adoption mobility between states [6]. Weighted event studies extend the synthetic control method (SCM) of [1] to the staggered adoption event study framework and cleanly nest within the fixed effects approaches of Eq. 1 and 2. To correct for imperfect pre-treatment balance, weighted event study augments a “partially pooled” SCM estimator with a fixed effects outcome model. Synthetic controls are constructed based on the balance of residualized pre-treatment outcomes; in this way, the approach builds upon recent research on doubly-robust estimators with an extension to the staggered adoption setting [2,3,6,14].^8^

While weighted event study design provides an appealing means for weakening the difference-in-differences identifying assumptions (softening to balance against a weighted combination of still-untreated states), it places additional requirements on the utilized data. In our context, synthetic control weights are obtained by matching treated units on residualized pre-treatment outcomes for the pre-mandate period of February 24 through March 18. Consequently, we require sufficient variation in these pre-treatment period outcomes to populate the donor pool and allow for construction of plausible control units. While the variation in travel patterns during this period is more than sufficient for obtaining successful synthetic control units for mobility analyses, the same cannot be said for health models. Table 2 showed that California’s statewide mandate on March 19 preempted much of the national health crisis, with on average under 1.5 days per state exhibiting variation in deaths for the pre-mandate period. As a result, we report weighted event study models for mobility analyses only, and limit health specifications to unweighted event studies.

We present results for both traditional unweighted event studies and weighted event studies in the form of traditional event study graphs. We report coefficient estimates for the event-time coefficients and 95% standard errors clustered at the state level for unweighted event studies, and plot 95% jackknife standard errors for weighted event studies [3].

## Results

Across the United States, COVID-19 upended daily routines. As a result of layoffs, revised work-from-home guidelines, school closures, family needs, and state policies, travel behavior changed dramatically in the U.S. from February through April 2020. Figure 1 plots over time the changes in average distance traveled (*AḊT*), visits to non-essential businesses (*NĖV*), and the unique human encounter rate (*EṄC*) per day for all U.S. states, measured as the percentage point change relative to typical pre-COVID-19 baseline levels. The solid line plots the average for the first four states to implement mandatory stay-at-home policies: California (implemented March 19), Illinois (March 21), New Jersey (March 21), and New York (March 22). The dotted line plots the average for the 15 states that adopted stay-at-home mandates later in the sample period, while the dashed line plots the daily average for the remaining 30 states that had yet to adopt a stay-at-home mandate by end-of-day March 25.^9^

**Figure 1:**
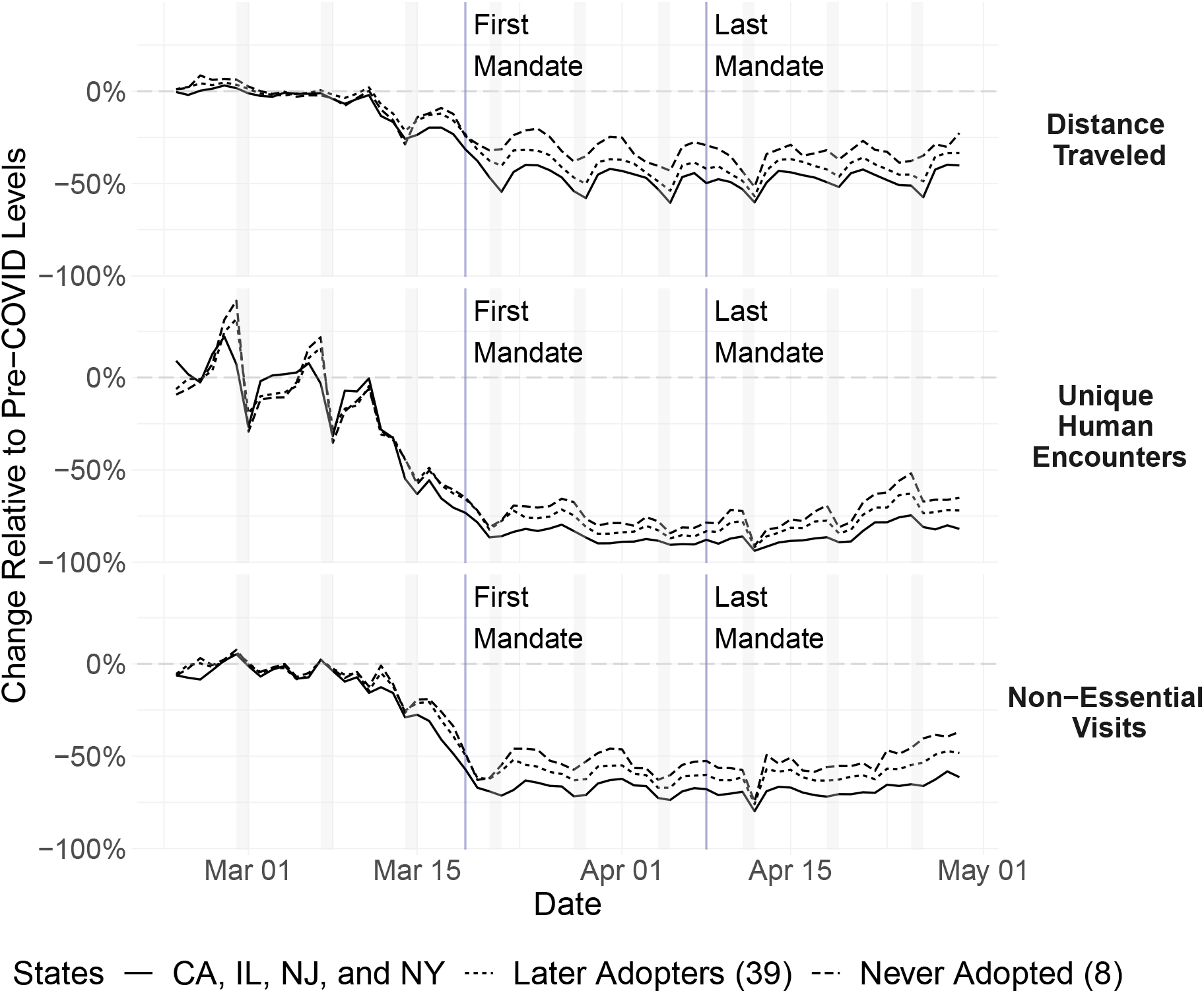
Changes in travel activity and social distancing. Each series represents the change in each day’s mobility measure relative to pre-COVID-19 levels for the given group of states. The solid line corresponds to the average change for the four states that implemented stay-at-home mandates by end-of-day March 22 (California, Illinois, New Jersey, and New York). The dotted line plots the average for the 39 states that adopted statewide mandates at later points, while the dashed line represents the average for the eight states that never adopted a statewide mandate. The first panel plots changes in average distance traveled, the second changes in unique human encounters per square kilometer, and the third changes in visits to non-essential businesses. The gray bars designate weekend days. The vertical lines indicate the dates of the first and last statewide stay-at-home mandates (March 19 and April 8).

Travel behavior in late February and through the first week of March looks largely typical for distances traveled and visits to non-essential businesses, with small fluctuations relative to baseline activity levels for all states. The change in the human encounter rate exhibits much greater variation throughout the week, increasing over the course of the work week before falling considerably over the weekend. Despite this greater within-week variation, the average human encounter rate for all states finishes the work week of March 2-6 above baseline levels.

Beginning the week of March 9, residents across the country began deviating from typical travel patterns. By Wednesday March 11, residents of all states had begun reducing their distances traveled, trips to non-essential businesses, and encounters with others relative to pre-COVID-19 norms. Initially, changes to mobility patterns in early-adoption states are largely indistinguishable from those for other states; by March 15, residents across all three groups had reduced travel distance by 8 to 13 percentage points, unique human encounters by 28 to 29 percentage points, and visits to non-essential visits by 12 to 17 percentage points.

By March 18, before the first statewide mandate went into effect, these declines had grown dramatically in magnitude. The change in travel distances fell further to between −12 and −23 percentage points, with changes between −34 to −49 percentage points for non-essential visits. Unique human encounters had already fallen between −61 and −71 percentage points relative to pre-COVID-19 baseline levels, a dramatic indicator of extensive social distancing occurring even before statewide orders required such behavior.

By the time many state implemented their policies in the coming weeks, travel behavior and social interactions had already largely bottomed out. While gaps between early adopter, later adopter, and never-adopter states increase slightly early in this period, they remain largely stable from the week of March 23 on, as do general within-week patterns.

Figure 1 provides initial evidence that changes in travel behavior are correlated with the decisions of whether and when to adopt stay-at-home mandates. Following the start of statewide mandate adoption on March 19, residents of early adopter states exhibit larger magnitude reductions every single day through April 29 across all three measures. Each week during this period, mean encounter rates in early adoption states are consistently 10 to 16 percentage points lower than in states that never adopted mandates, with a larger weekly gap in travel distance (between 13 and 19 percentage points) and a similar 12 to 22 percentage point gap for non-essential visits. Trends for later-adopting states fall between early and never-adopting states, with late adopters displaying declines between 5 and 7 percentage points larger in magnitude than never-adopters for human encounter rates, between 7 and 10 percentage points for travel distance, and between 6 and 10 percentage points for non-essential visits.

### Effect of Stay-at-Home Mandates on Daily Mobility and Social Distancing

Figure 1 provides preliminary evidence that residents across the country drastically reduced travel activity and engaged in extensive social distancing prior to the adoption of statewide mandates. We next present estimates of empirical models designed to identify any additional changes in mobility and social distancing patterns attributable to states’ stay-at-home mandates. We begin by presenting results of the staggered difference-in-differences treatment effect estimates in Table 3 before discussing the unweighted and weighted event study results in Figures 2 to 4.

**Table 3:** Statewide Stay-at-Home Mandates, Travel Activity, and Social Distancing

| | $\dot{A}DT$ | $\dot{A}DT$ | $\dot{N}EV$ | $\dot{E}NC$ | $\dot{A}DT$ | $\dot{A}DT$ | $\dot{N}EV$ | $\dot{E}NC$ |
| --- | --- | --- | --- | --- | --- | --- | --- | --- |
|  | (1) | (2) | (3) | (4) | (5) | (6) | (7) | (8) |
| $SAH_{it}$ | -4.454**<br>(2.074) | 2.495<br>(2.113) | 2.629<br>(1.927) | 3.050<br>(4.598) | -5.508***<br>(1.036) | -6.992***<br>(1.454) | -2.149**<br>(0.880) | -3.506***<br>(0.981) |
| $\bar{Y}$ | -26.59 | -29.29 | -36.23 | -53.57 | -26.59 | -29.29 | -36.23 | -53.57 |
| Pre-Period $\bar{Y}$ | -3.98 | -4.98 | -7.02 | -14.85 | -3.98 | -4.98 | -7.02 | -14.85 |
| State + Date FE | Yes | Yes | Yes | Yes | Yes | Yes | Yes | Yes |
| Cohort Pre-Trends | No | Yes | Yes | Yes | No | Yes | Yes | Yes |
| $N$ | 3,366 | 3,366 | 3,366 | 3,300 | 3,366 | 3,366 | 3,366 | 3,300 |
| Adjusted $R^2$ | 0.926 | 0.896 | 0.955 | 0.954 | 0.929 | 0.901 | 0.955 | 0.954 |

\* $p < 0.10$ , \*\* $p < 0.05$ , \*\*\* $p < 0.01$ . Standard errors are clustered at the state level. These models estimate the effect of statewide stay-at-home mandates on travel activity and social distancing. The dependent variables measure the percentage point changes in average distances traveled, visits to non-essential businesses, and unique human encounters for the same day of the week relative to the pre-COVID-19 baseline level (average of Feb 10 - Mar 8). A coefficient of one indicates a marginal effect of a 1 percentage point increase in travel relative to pre-COVID-19 levels, controlling for time and the average COVID-19 mobility change in the state during the sample period. *Cohort Pre-Trends* indicates that outcomes have been residualized of timing cohort pre-trends (following Goodman-Bacon 2018).

**Figure 2:**
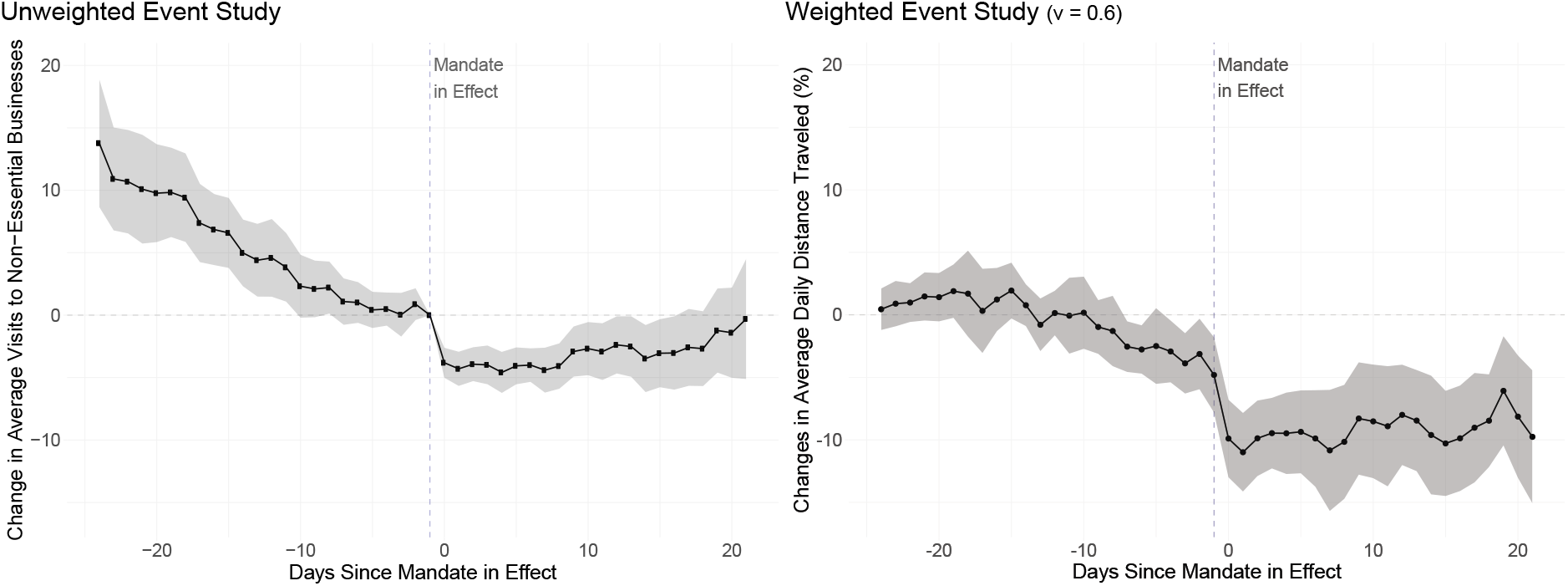
Unweighted and weighted event studies for changes in average distances traveled. The unweighted event study (left panel) plots regression coefficients for dummy variables equal to one for being *k* days away from the first effective date of each statewide stay-at-home mandate, with 95% confidence intervals represented in the gray band. A point estimate of −10 indicates a 10 percentage point greater decline in the average distance traveled per day for a state *k* days since mandate adoption relative to the day prior to mandate adoption (*k* = −1). The right panel plots equivalent point estimates and jackknife 95% confidence intervals from a weighted event study, with partially pooled synthetic controls constructed to match treated units on residualized pre-treatment outcomes with 60% of synthetic control weights obtained from pooled versus individual synthetic control weights (*ν* = 0.6).

**Figure 3:**
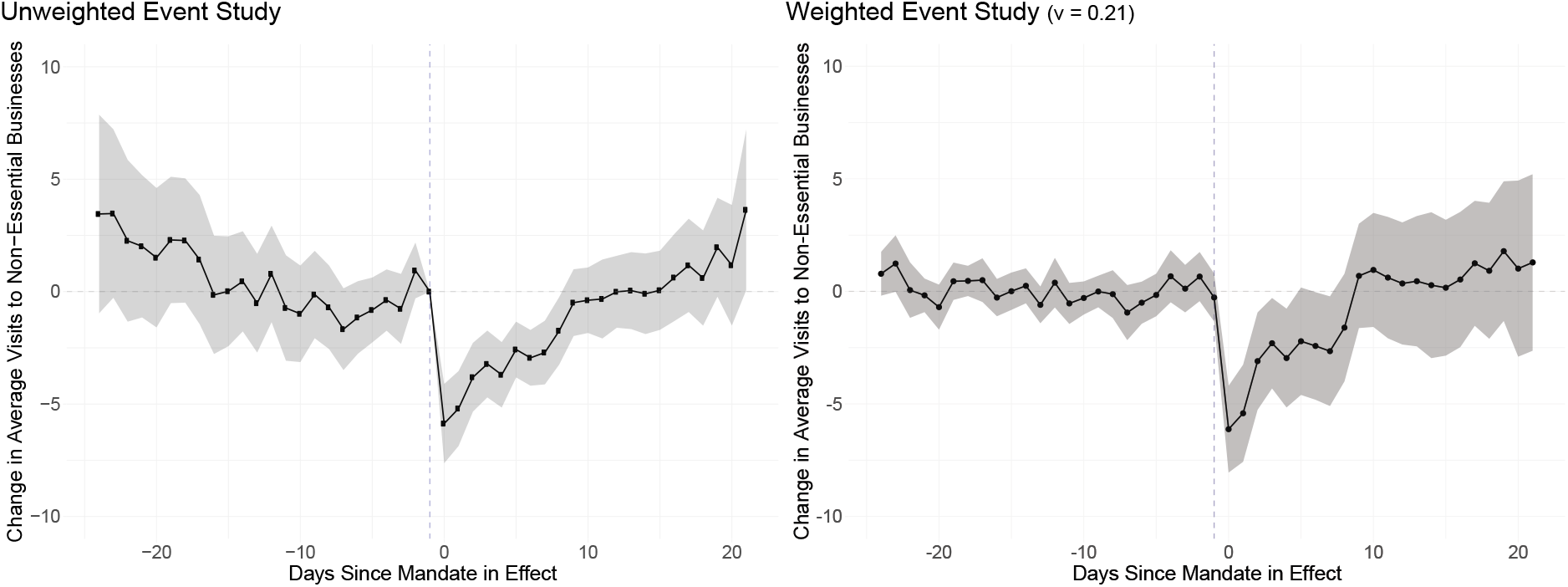
Unweighted and weighted event studies for changes in visits to non-essential businesses. The unweighted event study (left panel) plots regression coefficients for dummy variables equal to one for being *k* days away from the first effective date of each statewide stay-at-home mandate, with 95% confidence intervals represented in the gray band. A point estimate of −10 indicates a 10 percentage point greater decline in the average distance traveled per day for a state k days since mandate adoption relative to the day prior to mandate adoption (*k* = −1). The right panel plots equivalent point estimates and jackknife 95% confidence intervals from a weighted event study, with partially pooled synthetic controls constructed to match treated units on residualized pre-treatment outcomes with 21% of synthetic control weights obtained from pooled versus individual synthetic control weights (*ν* = 0.21).

**Figure 4:**
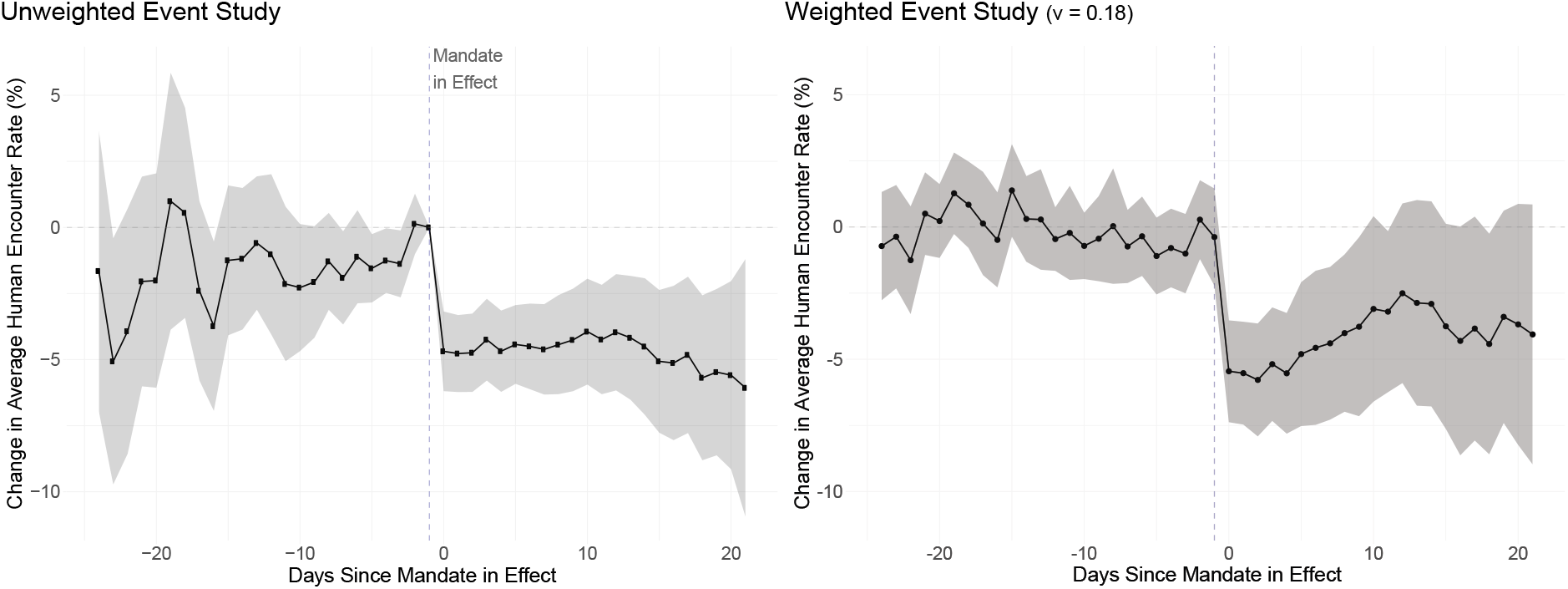
Unweighted and weighted event studies for changes in the unique human encounter rate. The unweighted event study (left panel) plots regression coefficients for dummy variables equal to one for being *k* days away from the first effective date of each statewide stay-at-home mandate, with 95% confidence intervals represented in the gray band. A point estimate of −10 indicates a 10 percentage point greater decline in the average distance traveled per day for a state k days since mandate adoption relative to the day prior to mandate adoption (*k* = −1). The right panel plots equivalent point estimates and jackknife 95% confidence intervals from a weighted event study, with partially pooled synthetic controls constructed to match treated units on residualized pre-treatment outcomes with 18% of synthetic control weights obtained from pooled versus individual synthetic control weights (*ν* = 0.18).

Table 3 presents difference-in-differences estimates from Eq 1 for the effect of stay-at-home mandates on travel activity across all three mobility measures. Columns (1) - (4) report coefficient estimates for the treatment effect restricted to only the first four adopters’ mandates (CA, IL, NJ, and NY) while columns (5) - (8) report estimates using variation for all 43 adopting areas to identify the treatment effect. Columns (1) and (5) report treatment effects with state and date fixed effects, while columns (2)-(4) and (6)-(8) also include dependent variables residualized of timing cohort pre-trends [19].

Comparing columns (1) and (2) illustrates the bias present when not controlling for cohort specific trends in the presence of dynamic adoption timing and treatment effects. In column (1) we estimate a −4.5 percentage point change in average distance traveled due to the first four states’ early mandates (for the sake of brevity we report equivalent models for *NĖV* and *EṄC* that yield similar magnitude estimates in Appendix C.) When we account for differences in adopters’ pre-trends across timing cohorts in column (2), the treatment effect estimate on *SAH_it_* changes sign and loses all statistical significance (the same is true for the other two measures of mobility). Once we account for these differences in pre-trends between early and later adopting cohorts, we fail to identify any differential effect of early adopters’ mandates across all measures in columns (2)-(4).

Columns (5) to (8) report equivalent estimates using adoption of all statewide mandates to identify the treatment effect estimate 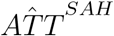. Here, with much greater variation in adoption timing, ATT estimates are identified both through comparisons of changes in treated units to changes in states that never adopted a mandate and through comparisons of states that had adopted and had yet to adopt at a given point in time. Comparing column (5) with column (6), we once again see the change in point estimates when accounting for cohort pre-trends. In this case, however, controlling for differences in pre-trends does not eliminate statistical significance and yields an ATT estimate statistically indistinguishable from that in column (5) (p = 0.15).

In our preferred specifications in columns (6) to (8), we estimate large treatment effects relative to pre-mandate behavior changes. Looking at estimates for changes in average distances traveled, we observe a treatment effect estimate of −6.99 percentage points for column (5), statistically significant beyond the 1% level. That is, once a statewide mandate is implemented, we estimate a 6.992 percentage point reduction in the change in average distance traveled relative to control states. This represents an additional 140% decline relative to the pre-mandate change of −4.98, and an additional 24% reduction relative to average changes over the entire sample period (−29.29). In column (7) we estimate a 2.149 percentage point reduction in visits to non-essential businesses per day relative to control states (a 31% and 6% additional reduction relative to pre-mandate and full sample average reductions, respectively). Turning next to changes in human encounter rates in column (8), we obtain an ATT estimate of a −3.506 percentage point decline per day after a mandate is implemented. Once again the treatment effects are statistically significant beyond the 1% level. This mandate effect corresponds to a 27% additional reduction relative to pre-mandate average changes of −14.85, and a 7% reduction relative to the −53.57 average change observed over the full sample period.

#### Decomposing the Difference-in-Differences ATT

A potential concern of the difference-in-differences estimator relates to the weighting of individual periods. Under staggered adoption, the estimated treatment effect can be expressed as a weighted average of all unique two-period by two-group difference-in-difference estimators [19]. Weights are implicitly assigned to each timing cohort and unit, proportional to the variance of the treatment indicator in each period and the size of each cross-sectional group. A key implication of these weights is a favoring of units treated near the middle of the sample period, with non-convexity indicating a potential for negative weights [2,8,16]. Another consequence is that negative treatment effects could also be obtained even when the effects of stay-at-home mandates for all adopting states are positive [12].

To shed light on the implicit weighting of the difference-in-differences ATT estimates presented in Table 3, we decompose the treatment effect estimates from columns (6) to (8) of Table 3 into their component two-by-two comparisons following [19]. We find that over half the overall difference-in-differences estimate’s weight is placed on comparisons of mandate states to never-adopter states (bottom panel). With adoption timing spanning March 19 to April 8, two-by-two comparisons can be made across many more cohorts and donor pools; in total, 18 comparisons are made between treatment cohorts and never-treated states, with 306 different comparisons between early and later adopters. More than half the overall ATT weight is given to comparisons of treatment cohorts versus pure control units, comprising 56-57% of the estimate across activity measures. The remaining weight is split evenly between comparisons of timing cohorts, with 21-22% of ATT weight given to comparisons of early treated units against later treated units still in the donor pool, and to later treated units post-treatment relative to previously-treated states. Except for early versus later treated units for average distances traveled, we observe consistently negative average ATT estimates across all three mobility measures, showing that treatment effect heterogeneity is primarily constrained to the size of reductions in travel activity. See Appendix B for a detailed presentation of the decomposition results and a more thorough discussion of the approach.

### Unweighted and Weighted Event Studies of Mobility and Social Distancing

To investigate the dynamic nature of mobility and social distancing responses to stay-at-home mandates and to address concerns regarding imbalances in changes to mobility patterns in the pre-mandate period, we next present results of unweighted and weighted event studies. Unweighted event studies avoid the implicit weighting concerns of the difference-in-differences estimator and allow an understanding of how treatment effects evolve and persist over time. As weighted event studies construct counterfactuals for each adopting state by balancing on pre-treatment outcomes, the obtained ATT estimate will more closely reflects the comparison of a given post-adoption period for each state to an appropriate trend from the pool of donor units available at each point in time.

Figures 2, 3, and 4 plot event study graphs for both unweighted and weighted event studies for average distances traveled, visits to non-essential businesses, and the human encounter rate. The left panel in each figure reports coefficient estimates and 95% confidence intervals for an unweighted event study estimated by Eq. 2 using outcomes residualized of cohort pre-trends. The right panel reports results from an equivalent weighted event study with the weight between separate and pooled synthetic control weights set equal to 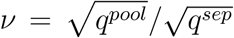, the ratio of the square roots of pooled to separate SCM imbalance [6].^10^ The x-axis of each plot reports event time, indicating the number of days elapsed since a state’s stay-at-home mandate entered into effect. An event time of zero indicates the first full day a state’s mandate was in effect. The unweighted event study relies on the parallel trends assumption required for the overall difference-in-differences approach, while the doubly-robust approach of the weighted event study necessarily imposes balance on changes in pre-treatment outcomes.

The unweighted event study (left panel) plots day-to-day ATT estimates averaged across all adopting states obtained from a two-way fixed effects control approach akin to columns (6), (7), and (8) in Table 3, with a vector of dummy variables for being each of *k* ∊ {−24, 21} days relative to mandate adoption. The day prior to adoption (*k* = −1) is normalized to zero, such that all point estimates are interpreted as a differential change in a given travel outcome on the *k^th^* day since mandate adoption relative to the day immediately preceding adoption. 95% confidence intervals clustered at the state level are reported in the gray band. Estimates statistically distinguishable from zero in the post-period measure the treatment effect of stay-at-home mandates on mobility patterns, decomposed by day. Non-zero estimates in the pre-mandate period (k < 0) are evidence that the difference-in-differences parallel trends assumption is likely violated: residents of adopting states were already differentially modifying their travel behavior relative to residents of control states prior to any statewide mandates requiring such behavior.

Turning first to changes in average distance traveled, Figure 2 shows the extent of pre-trends in the unweighted event study and the impact of internalizing these pre-trends in the weighted event study. In all periods 24 to 10 days prior to mandate adoption, adopting states display markedly higher average travel distances between 4 and 14 percentage points relative to control states. Pre-period point estimates remain positive but lose statistical significance for periods −9 to −2. Once a mandate is adopted, we observe an immediate reduction of 4 percentage points that persists for 9 days before gradually attenuating over the remaining post periods. The weighted event study with *ν =* 0.6 (60% of the weight given to the pooled synthetic control weights and 40% to the individual weights) in the right panel displays a greatly-improved overall pre-treatment match, with balance now achieved for periods −24 to −7 with slightly reduced fit in periods −6 to −2. We now observe persistence of the estimated mandate effect, with travel distance falling discontinuously immediately after a mandate and persisting below levels observed in the synthetic control units across all post-mandate periods. Averaging event day-specific ATT estimates across the entire mandate period yields magnitudes only slightly larger than those obtained through the staggered difference-in-differences approach in column (6) of Table 3, with an overall weighted event study ATT estimate of −8.97. This suggests that, while imbalance pre-trends may be present in the data, they impose only mild attenuation bias on the difference-in-differences estimates.

Looking next at models for changes in non-essential visits in Figure 3 and human encounter rates in 4, we see reduced evidence of differential trends in the unweighted event studies and more comparable weighted event studies as a result. Pre-treatment point estimates are statistically insignificant in all periods for non-essential visits and in 17 of 23 periods for human encounter rates. The unweighted event study for non-essential visits in the left panel of Figure 3 shows a 6 percentage point reduction the first day of mandate adoption that rapidly attenuates and becomes statistically insignificant after 8 days. The weighted event study in the right panel displays greater balance with narrow confidence intervals in all pre-treatment periods, with point estimates that are statistically indistinguishable from those in the unweighted event study (with slightly larger confidence intervals). The unweighted event study for human encounter rate changes in Figure 4 presents some evidence of differential trends, with an immediate and persistent treatment effect between −5.5 to −4.5 percentage points per day throughout the post-adoption period. Pre-treatment balance once again improves in the weighted event study, with treatment effect point estimates somewhat attenuated but largely indistinguishable from those obtained from the unweighted event study. Overall ATT estimates are once again in line with those obtained from the difference-in-differences models, with an overall mandate effect of −0.84 for non-essential visits (compared to −2.15 for column (7) of Table 3) and −4.14 for encounter rates (compared to −3.51 for column (8) of Table 3). These estimates are highly robust to the specific choice of *ν*; overall ATT estimates fall between −9.94 and −8.96 for average distance traveled, between −1.08 and −0.81 for non-essential visits, and between −4.17 and −3.51 for human encounter rates across the space of *ν €* [0, 1].^11^

The consistency of estimates between the difference-in-differences and both the unweighted and weighted event study approaches provides confirming evidence that statewide stay-at-home mandates elicited further reductions in travel activity by affected residents. Event studies provide additional detail as to how these responses evolved, showing that reductions occurred immediately upon policy implementation and largely persisted even as residents were subject to the policies for three full weeks. This pattern is especially true for average distance traveled and human encounters, suggesting that residents of mandate states continued to socially distance, a key avenue for reducing the potential transmission of COVID-19.

#### Take Away and Additional Evidence on Mobility

Taken together, Figure 1 and the subsequent analyses reveal two main facts. First, that people across the U.S. decreased their travel and rate of human encounters early in the pandemic, preempting statewide requirements. Second, we find that statewide stay-at-home mandates are related to additional reductions in all our measures of travel activity. Distance traveled is positively linked to an increased number of social trips across all modes of transportation, suggesting that the observed decreases likely reflect a decline in unique trips away from home as well. As travel activity is a main source of social interaction beyond one’s immediate family [32] and travel to non-work locations increases the probability of co-location with others, these reductions in distances traveled likely reflect commensurate decreases in physical interactions with those outside of one’s immediate family. Our estimates for changes in unique human encounters reflect these previous studies, providing evidence of further social distancing once states adopted a stay-at-home mandate. Further, these findings are not limited to the Unacast mobility measures; use of Google’s COVID-19 Community Mobility Reports estimates similarly large and statistically significant effects of statewide mandates.^12^ All this provides consistent, preliminary evidence that stay-at-home mandates are having the intended effect of inducing greater social distancing than would occur otherwise, helping to reduce the opportunities for communication of COVID-19 within communities.

### Effect of Stay-at-Home Mandates on Health Outcomes

Figure 5 plots over time the average daily death rates per 100,000 residents.^13^ As in Figure 1, the solid line plots the average for the first four states to implement mandatory stay-at-home policies, while the dotted line plots the average for later adopters of statewide mandates, and the dashed line plots the daily average for never-adopter states.

**Figure 5:**
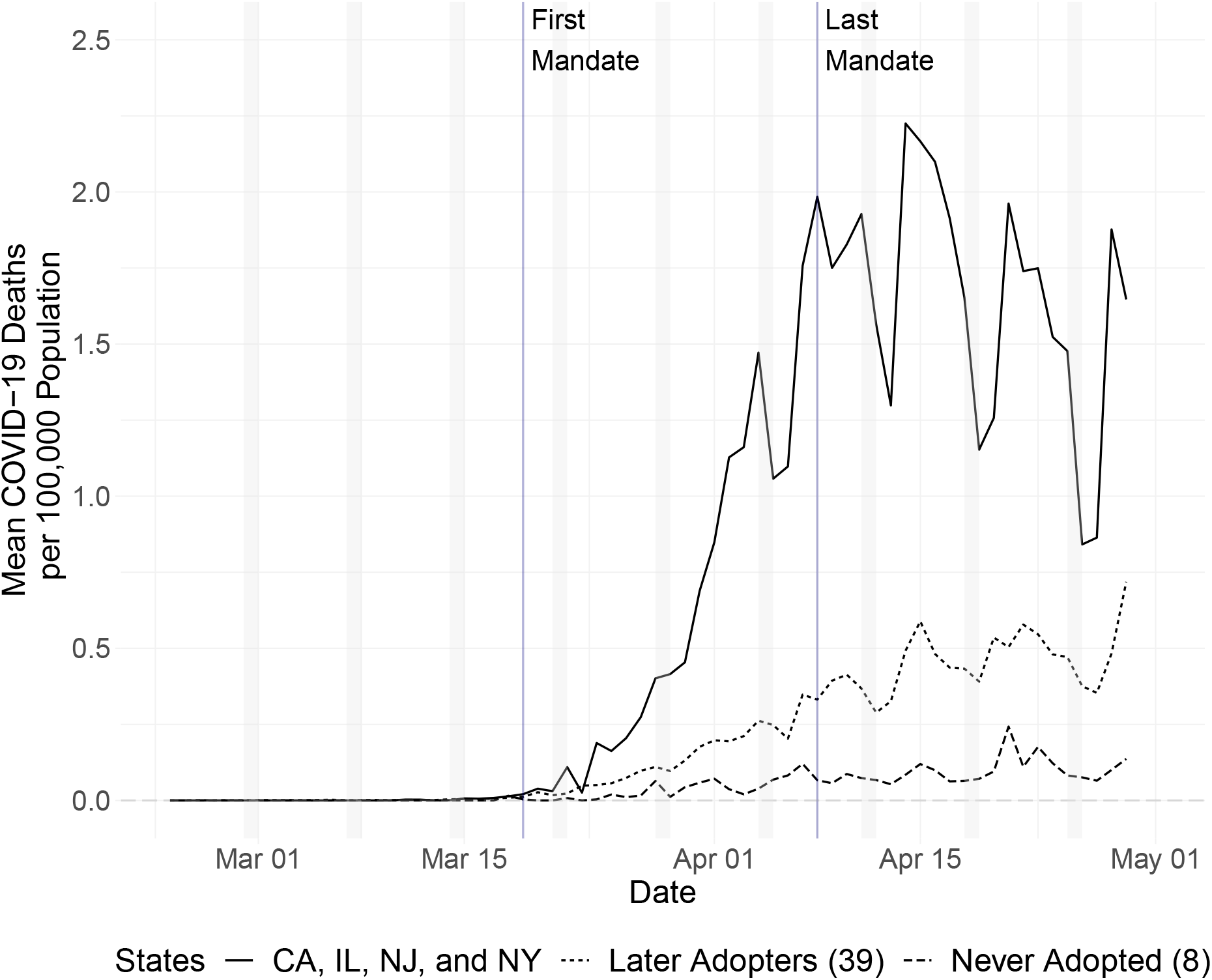
Each series represents the day’s death rate (per 100,000 residents) for the given group of states. The solid line corresponds to the average change for the four states that implemented stay-at-home mandates by end-of-day March 22 (California, Illinois, New Jersey, and New York). The dotted line plots the average for the 39 states that adopted statewide mandates at later points, while the dashed line represents the average for the eight states that never adopted a statewide mandate. The gray bars designate weekend days. TThe gray bars designate weekend days. The vertical lines indicate the dates of the first and last statewide stay-at-home mandates (March 19 and April 8).

The lack of variation in death rates prior to the mandate adoption period is immediately evident. As shown in Table 2, fewer than half of states had experienced a COVID-19 death by March 18, with average death rates under 0.002 per 100,000 population for the pre-mandate period. Differences begin manifesting across the three groups of states the week following the start of the adoption period, with death rates growing at a much faster rate for early adopter states relative to the other two groups. By April 8, average death rates in early adopter states were roughly five times those of later adopters, with never-adopters displaying rates less than half those of later adopters.

Figure 5 alludes to many of the empirical challenges facing successful identification of the causal impacts of stay-at-home mandates on COVID-19 mortality and morbidity. As most states adopted policies relatively early in their region’s outbreak, caseloads and death rates in states just adopting statewide stay-at-home mandates often look much different in comparison to earlier adopters. Further, as transmission of COVID-19 can take weeks before symptomatic cases, hospitalizations, and deaths occur, it likely would take weeks or even months before death rates reflect the impacts of extensive changes to social distancing behavior due to statewide policies. As a result, observed health outcomes can reasonably be expected to worsen shortly after mandate adoption – indicating that naive estimation strategies would likely indicate that stay-at-home mandates and additional social distancing *causes* more COVID-19 deaths. The lack of variation in death rates prior to the adoption of statewide policies also prevents the use of matching techniques like weighted event studies that could account for differences in the stage of pandemic in treated versus untreated states. Finally, death rates in never-adopter states were dramatically lower than in adoption states throughout the adoption and post-adoption periods, indicating their inclusion as control units would likely accentuate the pre-existing identification issues.

To combat the empirical challenges above, we evaluate the mortality data using differences-in-differences, as well as a standard event study model. This also allows us to account for the changes in mobility behavior before the stay-at-home mandates. To the extent that differences in these mobility behavior pre-trends exist between states, and that health outcomes change in response to changes in travel activity and social distancing, preferred models must adequately control for these prior disparities.

#### Difference-in-Differences Estimation of Health Outcomes

Table 4 presents estimates from Eq 1 for the effect of stay-at-home mandates on death and hospitalization rates using the staggered difference-in-differences estimator. Columns (1) and (2) report estimates for death rates per 100,000 population while columns (3) and (4) report equivalent estimates for hospitalization rates. The dependent variable in both columns is the residual death rate after removing the predicted pre-trend death rate for each mandate timing cohort [19].^14^ All models include state and date fixed effects. To account for differences in pre-mandate mobility pattern changes that may have affected subsequent health outcomes, all columns include interactions of changes in all three mobility measures with an indicator for the pre-adoption period (equal to zero on or after March 19 for death models, and zero on or after the date of mandate adoption for hospitalization models).

**Table 4:** Stay-at-Home Mandates and COVID-19 Mortality and Morbidity

|  | Death Rates |  | Hospitalization Rates |  |
| --- | --- | --- | --- | --- |
|  | (1) | (2) | (3) | (4) |
| $SAH_{it}$ | -0.169*<br>(0.096) | -0.131*<br>(0.068) | -5.597**<br>(2.654) | -6.038**<br>(2.777) |
| $\dot{ADT} \times \text{Pre-Period}$ | 0.005<br>(0.007) | 0.010<br>(0.007) | 0.034*<br>(0.018) | 0.035*<br>(0.019) |
| $\dot{VIS} \times \text{Pre-Period}$ | 0.026*<br>(0.014) | 0.016**<br>(0.006) | 0.011<br>(0.018) | 0.007<br>(0.020) |
| $\dot{ENC} \times \text{Pre-Period}$ | 0.006<br>(0.007) | -0.001<br>(0.001) | 0.050*<br>(0.026) | 0.056*<br>(0.028) |
| Pre-Period $\bar{Y}$ | 0.0016 | 0.0007 | NA | NA |
| $\bar{Y}$ After First Death | 0.348 | 0.384 | 1.074 | 1.219 |
| Post-Period $\bar{Y}$ | 0.494 | 0.565 | 1.090 | 1.248 |
| Treatment Group | All | All | All | All |
| State + Date FE | Yes | Yes | Yes | Yes |
| Cohort Pre-Trends | Yes | Yes | No | No |
| Sample | 2wk Pre | SAH | 2wk Pre | SAH |
| $N$ | 2,245 | 2,772 | 1,084 | 916 |
| Adjusted $R^2$ | 0.538 | 0.470 | 0.598 | 0.589 |

\* $p < 0.10$ , \*\* $p < 0.05$ , \*\*\* $p < 0.01$ . These models estimate the effect of all statewide stay-at-home mandates on COVID-19 death and hospitalization rates per 100,000 population. *Cohort Pre-Trends* indicate outcomes de-trended by treatment cohort-specific linear trends (Goodman-Bacon 2018). All models control for $\dot{ADT}$ , $\dot{NEV}$ , and $\dot{ENC}$ prior to the first statewide mandate on March 19 for all states in death models and each state's mandate for hospitalization models. *2wk Pre* indicates sample trimmed to 2 weeks before each states' mandate and date of first COVID-19 death for never-mandate states, while *SAH* limits the sample to the states that ever adopt statewide mandates (42 and D.C.).

To focus comparisons on when and where outbreaks occurred, we estimate models using two subsets of the sample data. First, in columns (1) and (3) we trim observations for each state to the date of first death for never-adopting states and within two weeks of mandate adoption for the 43 states and D.C. that adopt mandates.^15^ In columns (2) and (4) we omit observations for states that never adopt mandates, narrowing the comparison group to states with similar outbreak trajectories and relying on variation in treatment timing within adopting states to identify the treatment effect.^16^

Across specifications, we find preliminary evidence that adoption of statewide stay-at-home mandates attenuated the mortality and morbidity consequences of COVID-19. Following adoption of a statewide mandate, we observe a decrease in daily death rates of between 0.13-0.17 fewer deaths per 100,000 population that is statistically significant at the 10% level when controlling for pre-period changes in mobility patterns. These effects are large relative to average death rates over the entire utilized sample periods: estimated post-mandate death rates fall by 34-49% compared to average death rates of 0.348-0.384 for periods after each state experienced its first death, 3 daily deaths per 100,000) and by 23-34% of the post-adoption period average rates (0.494-0.565 per 100,000 for the post-adoption period of April 9-29).

Treatment effect estimates for hospitalization rates are much larger in absolute and relative magnitudes. In columns (3) and (4) we estimate a decline of between 5.6 and 6.0 daily hospitalizations per 100,000 population. Compared to the full sample period average hospitalization rates of 1.05 and 1.17, these estimates suggest that, absent adoption of a statewide mandate, hospitalization rates would have been roughly five times greater than observed after each state’s first death (1.074-1.219 daily hospitalizations per 100,000) and in the post-adoption period (1.090-1.248 per 100,000). Mandate effect estimates are statistically indistinguishable from each other across samples for both death and hospitalization rates, supporting the notion that comparisons within treated states are driving the treatment effect.

Scaling the daily estimated mandate effects to the duration of a stay-at-home mandate suggests the importance of these policies in curbing the spread of COVID-19. For a 61 day mandate, this corresponds to 8.0-10.3 fewer deaths and 341-368 fewer hospitalizations per 100,000 population. For the average adopting state with a population of 6.8 million, our models predicts 543-701 averted deaths and 23,216-25,046 averted hospitalizations due to an implementaton of one two-month stay-at-home mandate. Across 43 adopting states, this represents 23,366-30,144 fewer deaths and roughly one million averted hospitalizations. By April 29, 55,478 deaths had occurred across all adopting states, suggesting that total deaths could have been 42-54% greater had states not adopted statewide stay-at-home mandates.

Across models we similarly observe signs that changes in mobility patterns prior to adoption of statewide mandates affected subsequent health outcomes. In models of death rates we obtain positive point estimates on 5 of 6 pre-period mobility controls, with estimates for non-essential visits achieving statistical significance at the 5-10% levels.^17^ As more trips to non-essential businesses increase human-to-human contact and potential exposure to infected individuals, we would expect a positive relationship between additional trips and worse health outcomes in later weeks. The point estimates of 0.016-0.026 are relatively large relative to pre-period changes in mobility patterns. Scaling effects by the average pre-period change in non-essential visits of −8.8 for mandate states, this represents a decline in average daily death rates between 0.141-0.229 deaths per 100,000 population – effects comparable to those of the stay-at-home mandates themselves.

Turning to hospitalization models, we find similar evidence of the effect of pre-period mobility pattern changes. We obtain positive point estimates on all mobility coefficients in columns (3) and (4), with statistical significance at the 10% level attained for coefficients on changes in average distances traveled and human encounter rates. Given the pre-period average changes of −4.98 for distance traveled and −14.85 for unique human encounters, these once again provide initial evidence that health outcomes improved in states where residents stayed at home and socially distanced early in the COVID-19 pandemic. Decomposing the difference-in-difference estimators provides additional support for this notion, as seen in Appendix B.

#### Mortality Event Studies

To investigate the dynamic nature of COVID-19 mortality responses to statewide stay-at-home mandates, we next report estimates obtained from unweighted event studies following Eq. 2.^18^ Figure 6 plots coefficient estimates and 95% confidence intervals for event times −14 through 21 for two different specifications. In the left panel we report estimates obtained from a model equivalent to column (1) of Table 4, where we use a 2-week pre-treatment sample and include controls for changes to pre-period mobility patterns. The right panel reports corresponding estimates when the sample is further limited to stay-at-home states only.^19^

**Figure 6:**
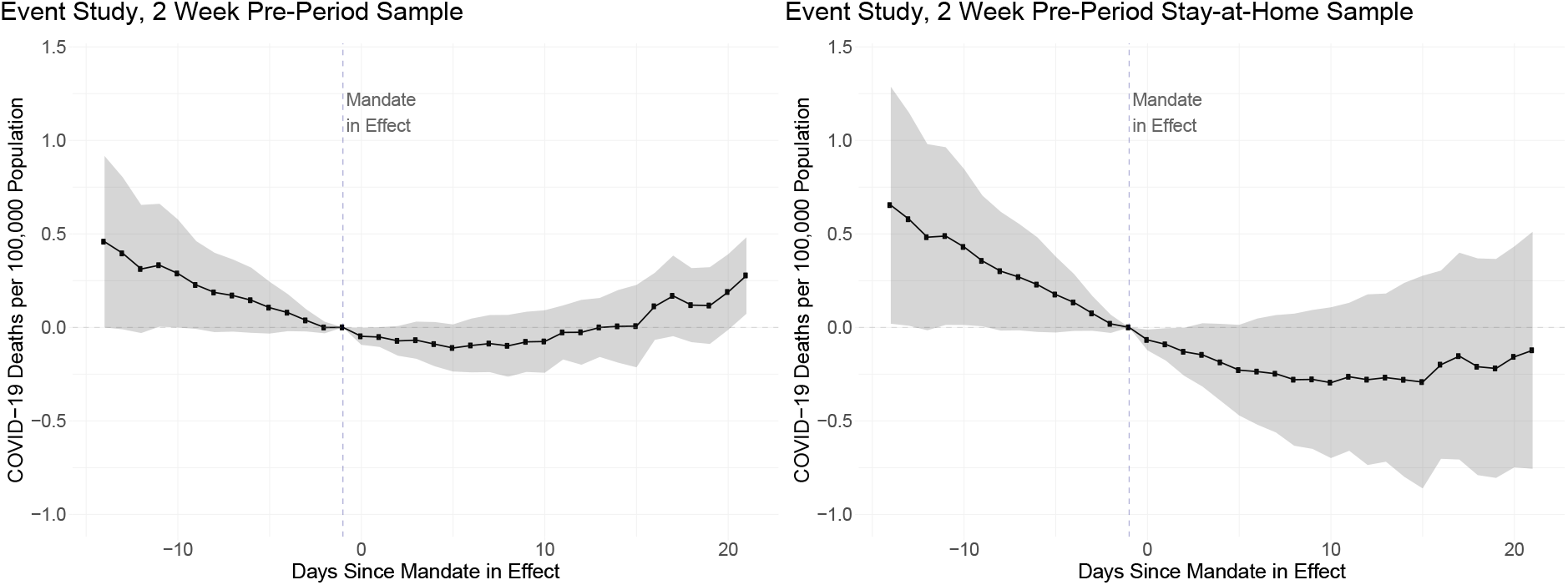
Unweighted event studies for death rates following statewide stay-at-home mandates. The unweighted event study in both panels plot regression coefficients for dummy variables equal to one for being k days away from the first effective date of each statewide stay-at-home mandate, with 95% confidence intervals represented in the gray bands. A point estimate of −10 indicates a 10 percentage point greater decline in the average distance traveled per day for a state k days since mandate adoption relative to the day prior to mandate adoption (*k* = −1). The left panel uses pre-treatment data trimmed to two weeks prior to each state’s stay-at-home mandate, and the date of first death for never-adopters (N = 2,147). The right panel further limits the sample to only states that ever adopt a statewide mandate (n = 2,102).

The lack of deaths in the pre-adoption period prevents estimation of a weighted event study, and unweighted event studies do suggest that trends in death rates differed for adopting and control states in the days prior to adoption. While coefficient estimates are statistically insignificant for all 13 estimated pre-treatment periods in the left panel and in 8 of 13 periods for the adopting state sample, we observe point estimates suggestive of large differences in pre-treatment trends. Once mandates come into effect, treatment effect estimates fall below zero. These negative point estimates continue for 10 days in the 2 week pre-treatment sample and throughout the post-treatment period for the stay-at-home sample, but all coefficients are statistically insignificant. From day 10 on, point estimates rise in the 2 week pre-treatment sample, only becoming statistically significant for the binned endpoint at event time 21.

Overall, the event studies provide little insight into the dynamics of stay-at-home mandates’ impacts on COVID-19 mortality. This is unsurprising, given the low statistical significance in the difference-in-differences models and the previously-discussed empirical challenges in this setting. Future work can focus on implementing additional techniques that can produce improved comparisons and achieve better pretreatment balance.

## Conclusion

Temporarily closing non-essential businesses and mandating that residents stay at home except for essential activity is the prime policy instrument currently employed by states to promote social distancing and slow the transmission of COVID-19. If effective, these policies will have reduced strain on the medical system and provided much-needed time for the development of pharmaceutical treatments that can reduce transmission rates and end the pandemic. If unsuccessful, states will have incurred large economic costs with few lives saved. Whether these mandates cause people to stay at home and engage in social distancing is a key requirement of a successful policy. Knowing whether such policies will have their intended effect is of increasing policy relevance, as all but eight states eventually adopted such policies. Understanding whether and how individuals reduce travel activity and maintain social distance in response to stay-at-home mandates is the primary empirical question we tackle in this paper.

First, we find that, by the time the average adopter had implemented its statewide mandate, residents had already reduced travel by considerable amounts relative to pre-COVID-19 levels. Average travel distances had already fallen by 16 percentage points, human encounter rates by 63 percentage points, and non-essential visits by 39 percentage points before the first statewide mandate came into effect, providing evidence of extensive social distancing occurring even before this was required by statewide orders.

Second, we find evidence that adoption of state-level stay-at-home mandates induced further reductions in all three travel activity measures. The staggered difference-in-differences models estimate a reduction in average distance traveled of 5.51 percentage points, a decline in visits to non-essential businesses of 2.15 percentage points, and a decrease in the rate of unique human encounters of 3.51 percentage points relative to pre-COVID-19 baselines. Estimated magnitudes remain highly comparable when directly accounting for differences in pre-mandate behavior for treatment and control states. Through the weighted event studies that construct control units to balance pretreatment travel behavior net of state fixed effects, we find large, statistically significant drops immediately following mandate implementation across all measures that persist for the duration of the sample period for distances traveled and human encounters. Resulting estimates of the overall mandate effects mirror those obtained from the difference-in-differences models, and are similarly sized for any mix of pooled and separate synthetic control weights.

Our estimates suggest that residents subject to stay-at-home mandates on average responded as desired to curb the spread of COVID-19. Our empirical approaches isolate the mandate effect from other drivers of daily changes in travel activity levels and from pre-existing trends, and control for a host of potential confounding factors that differ between states that adopted policies relative to other states and those yet to adopt policies. Under these rigorous control approaches, we find persistent evidence of state mandates inducing further reductions in travel activity even after considerable earlier declines around the country. Further, our estimates are average treatment effects in response to statewide mandates only. Given the extent of prior school closures, new work from home abilities, and county-level stay-at-home policies, our findings represent only a portion of the way individuals responded to COVID-19 policies. As a result, our estimates represent a considerable lower bound on individuals’ comprehensive responses to all COVID-19 policies.

Finally, we provide initial evidence of the link between stay-at-home mandates and attenuated health impacts from COVID-19. Our findings suggest that stay-at-home mandates likely decreased daily deaths by 0.13-0.16 and daily hospitalizations by 5.6-6 per 100,000 population in the average mandate state. Further, we observe that changes to mobility patterns prior to the adoption of statewide mandates played a similar magnitude role in averting COVID-19 deaths. Projecting these findings to stay-at-home mandates spanning March and April in all adopting states, this corresponds with 23-30,000 averted deaths due to mandate-induced behavioral responses and 48-71,000 averted deaths across all behavior changes for mandate states in these two months. Our estimates indicate that, absent these policies, U.S. deaths from COVID-19 could have been 1.86-2.27 times higher than they actually were during March and April. Our results provide preliminary evidence that the changes in social distancing and travel behavior induced by statewide mandates likely played an important role in flattening COVID-19 epidemic curves in adopting states.

Our findings have important policy implications for the fight against COVID-19. First, individuals on average responded as intended to statewide mandates. Despite considerable prior reductions, residents heeded their states’ directives and stayed at home. Second, the declines in economic activity directly attributable to statewide mandates may be much smaller than previously thought. Because individuals around the country had already more than halved the quantity of trips taken to non-essential retail and service businesses, much of the lost business and resulting unemployment would have likely still occurred even if states had not adopted their stay-at-home policies. Further, as the mandate-induced reductions in visits to non-essential businesses amount to only one-tenth of the overall reductions since COVID-19, it is likely that loosening or removing statewide policies will not be sufficient to induce mobility patterns to quickly return to pre-COVID-19 levels. Further policies will be needed to ensure that individuals can safely resume activity and return to local businesses.

Our estimates do not take into account the benefits from avoided hospitalizations and other indirect health benefits from reduced travel activity and social distancing. Because reductions in travel distance and increased social distance likely decrease exposure to other potentially deadly illnesses, this is likely an underestimation of the overall health benefit of these policies. Further, the patterns in under-reporting and under-counting of COVID-19 deaths suggest that we likely underestimate the direct benefits of these policies. Future identification of additional COVID-19 deaths may prove difficult, as many death certificates list only the immediate cause of death and fail to report underlying diseases – likely understating the presence of COVID-19 [26]. Further, procedures for counting COVID-19 deaths may be correlated with adoption of stay-at-home mandates. If adoption of a state-level ordinance indicates additional preparedness on the part of the adopting state, then states that were slower to (or have yet to) pass stay-at-home mandates may also have been slower to properly attribute deaths to COVID-19, resulting in our estimated effects being understatements of the true effect. Given the challenges to proper identification of COVID-19 deaths, we may not know the true death count for years, or ever. We support continued efforts to obtain accurate counts of the mortality and morbidity consequences from COVID-19 to help ensure future research can provide sufficient policy guidance in the case of future pandemics.

## Data Availability

Data used for this analysis are from Unacast, Health Data are from the COVID 19 Tracking project, and policy data from The New York Times, and COVID-19 US state policy databases.

## Footnotes

1 See Table 1 in Appendix A for a complete list of all retail and service categories classified as non-essential.

2 For data quality reasons, we drop observations for Washington D.C. from our analysis of human encounter rates.

3 In contrast to *AḊT* and *NĖV*, which are normalized by the state’s day-of-week average from the entire pre-COVID-19 baseline period of February 10 through March 8, due to data limitations *EṄC* is only normalized using the state’s day of week average for February 24 through March 8. For more discussion, see Section 1.4 of Appendix A.

4 While the pre-COVID-19 baseline period used for normalization extends back to February 10, our state-by-day mobility panel only begins on February 24 and does not include daily observations for February 10-23. However, Figure 1 shows that travel patterns were largely indistinguishable from baseline levels prior to early March, providing evidence that behavior had not been substantially modified during the month of February.

5 We do not have data on the entire pre covid baseline period that was used to construct the measures we obtain from Unacast. We only have daily data for the baseline period from February 24th to March 8.

6 See the data Appendix A for all state-specific correlations between the Unacast measures and the retail and recreation measures from Google’s COVID-19 Community Mobility Reports.

7 See the Data Appendix A for tables correlating CVT, the New York Times, and Johns Hopkins’ death rates. This provides evidence for the insensitivity of our health findings to our choice of data source.

8 For each treated unit *k*, 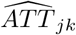 can be thought of as a doubly-weighted difference in differences estimator, wherein the change in the treatment unit *j* is obtained as the difference between the treatment unit’s outcome *k* periods post-adoption and its pre-period average, and the change in the control group is the average for equivalent changes for all donor units, weighted by partially pooled synthetic control weights. Averaging 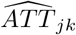 across all treated units at a given point in event time yields a period-specific treatment effect 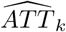 that can be thought of as equivalent to the dynamic ATT obtained from an unweighted event study design. Standard errors are obtained using a jackknife approach [3].

9 Colorado, Connecticut, Delaware, Hawaii, Idaho, Indiana, Louisiana, Michigan, New Mexico, Ohio, Oregon, Vermont, Washington, West Virginia, and Wisconsin all implemented similar stay-athome mandates between March 23 and March 26. Massachusetts adopted a stay-at-home advisory, recommending but not requiring that residents stay home.

10 See Appendix D for overall ATT estimates obtained across the space of *ν*. While an interior nu of 0:01 − 0:99 offers substantial imbalance reductions relative to the pooled or separate SCM cases, the optimal choice of *ν* is not immediately obvious. Estimating weighted event studies over the range of *ν* allows us to better understand how sensitive the overall ATT estimate is to the shift in weight from separate SCM for each state to a purely pooled SCM approach.

11 See Appendix D for plots of ATT estimates for each 0.01 value of *ν*.

12 See Appendix F for the complete complementary Google mobility analysis, replicating the same methodology controlling for state-specific flexible trends, state, and day fixed effects.

13 See Appendix A for a corresponding figure for hospitalization rates.

14 Not accounting for cohort pre-trends in death rates has no impact on estimates, which follows from the lack of COVID-19 deaths in the pre-mandate period. Only 25 states had experienced a COVID-19 death by March 19 and only 4 states had witnessed 10 or more deaths by that date (CA, GA, NY, WA). A similar technique cannot be done for hospitalization rates due to the lack of data in the pre-mandate period.

15 Washington is the only state in which COVID-19 deaths occurred more than 2 weeks prior to mandate adoption. Coefficient estimates are identical when observations are included for Washington beginning on February 27, the date of first death.

16 Results of alternative specifications including using the entire pre-mandate period are presented in Appendix E.

17 The lack of statistical significance across all three measures is not surprising given the level of collinearity between the measures. Across the entire sample we observe corr(*AḊT*, *VİS*) = 0.935, corr(*AḊT*, *EṄC*) = 0.875, and corr(*EṄC*, *VİS*) = 0.929.

18 We are unable to present equivalent estimates for COVID-19 hospitalization rates due to the lack of reporting prior to states’ mandates and throughout the entire pre-mandate period.

19 Results of alternative event study specifications are presented in Appendix E.

